# Speech Timing Measures Are Not Interchangeable Across Elicitation Methods in Senior Living Residents

**DOI:** 10.64898/2026.09.23.26363840

**Authors:** Kevin Mekulu, Nicole Etter, Timothy Brearly, Hui Yang

## Abstract

Speech-based cognitive monitoring for Alzheimer’s employs various tasks, but whether timing measures transfer across tasks is unclear. Twelve senior living residents completed four 60-second tasks: category fluency, two picture descriptions, and a self-generated narrative. Speech timing measures were calculated identically across tasks. Speech-rate correspondence among picture descriptions and narrative was substantial (mean ρ = 0.72). Correspondence with category fluency increased from Cookie Theft to contemporary picture to narrative in terms of speech rate (ρ = 0.36, 0.51, 0.64) and pause time ratio (ρ = 0.14, 0.45, 0.71). Measures were not interchangeable; correspondence followed an ordering consistent with shared retrieval demand.

## Introduction

Speech timing measures derived from briefly recorded tasks are often investigated as low-burden indicators of cognitive status in older adults [1-4]. Measures such as speech rate and pause duration can be extracted automatically from short samples without specialized equipment and impose minimal burden on the person being assessed. However, studies use markedly variable elicitation methods, including category fluency, picture description, and narrative tasks [5-7].

Elicitation methods impose different retrieval and discourse demands. Category fluency requires continuous retrieval from a semantic category without external content support. Picture description provides visual content that constrains discourse and cues lexical retrieval, often with administration-related pauses in stimulus provision. Self-generated narrative requires generating and organizing content without visual stimuli or categorical boundaries, although familiar autobiographical material may provide support. Timing measures may therefore correspond more strongly between tasks that share retrieval demands.

Direct comparisons are uncommon. Clarke et al. found that linguistic features and classification performance varied across five connected-speech tasks completed by the same participants [8] but did not examine within-person correspondence of identical timing measures. Other multitask protocols have combined task-derived features for classification rather than testing whether a participant’s relative standing on a measure is preserved across tasks [5].

We elicited four 60-second spoken-language samples per participant and applied identical definitions of speech rate, word count, pause time ratio, and pause-adjusted rate. We assessed cross-task correspondence and whether its ordering was consistent with differences in retrieval demand. The aim was to inform elicitation-task selection for speech-based monitoring protocols rather than establish criterion validity.

## Methods

### Participants and setting

Twelve residents from a continuing care retirement community in central Pennsylvania participated. Mean age was 85 years (SD = 3.2, range 80 to 92); seven were female. Four held graduate degrees, six held undergraduate or equivalent degrees, and educational attainment was unknown for two. Recordings occurred onsite in a quiet, private room during routine facility operations. Figure 1 summarizes the workflow.

**Figure 1.**
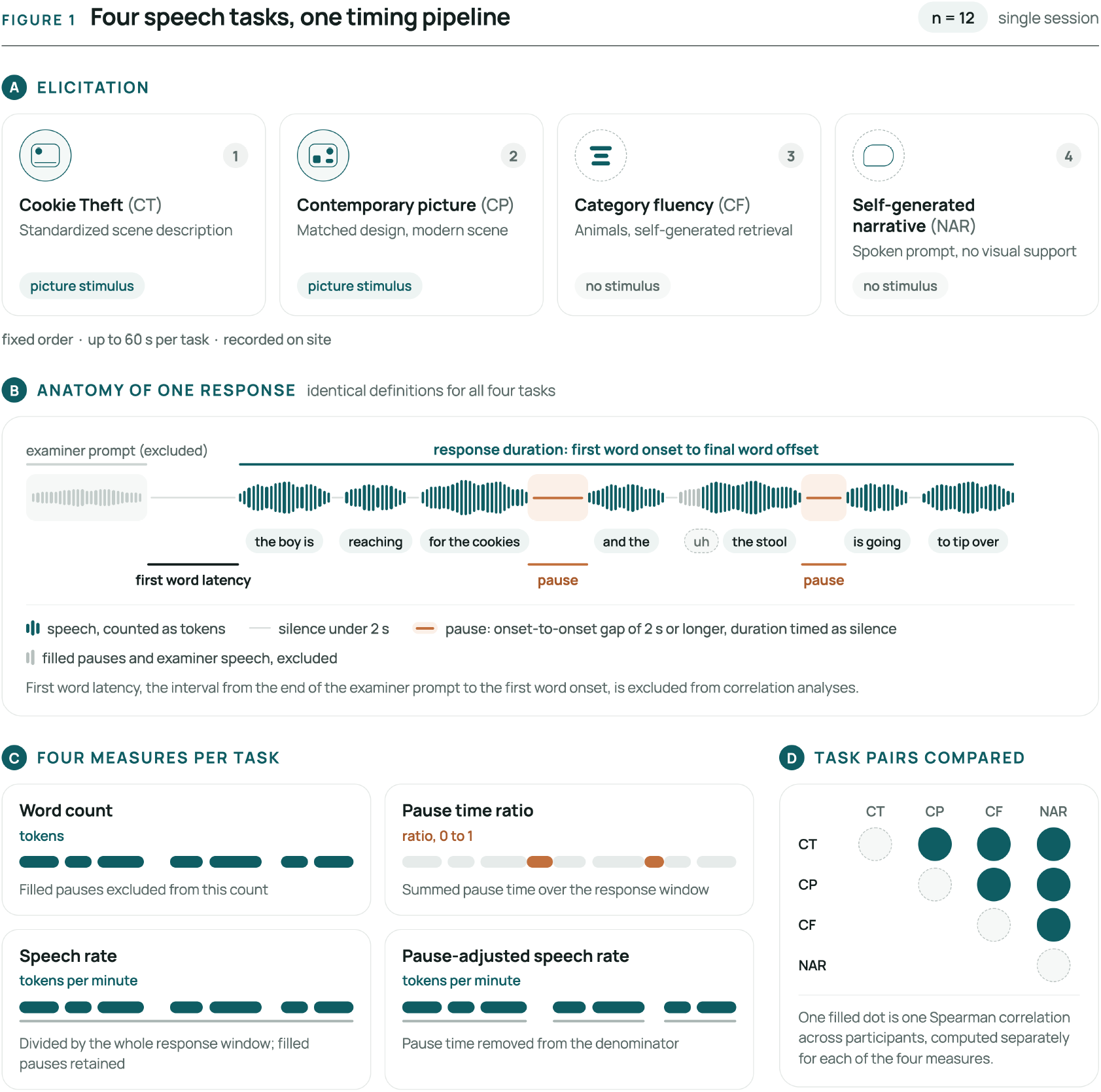
Study workflow. Twelve participants completed four elicitation tasks in one fixed-order session (A). Identical timing definitions were applied across tasks (B), yielding four speech timing and productivity measures per task (C). Spearman correlations were computed for all six task pairs separately for each measure (D). Filled pauses were retained in speech rate but excluded from word count and pause detection; examiner speech was excluded throughout.

### Speech sample stimuli

Four spoken-language samples were elicited from each participant, each allowing up to 60 seconds, in fixed order: Cookie Theft picture description, contemporary picture description, category fluency, and self-generated narrative. The order reflected sequencing from established to exploratory stimuli and was not designed to manipulate demand. The two picture descriptions and narrative were classified as connected-speech samples because they elicited continuous multiword utterances; category fluency elicited isolated category exemplars.

The contemporary image was designed as a modern analogue of Cookie Theft, with multiple actors, concurrent activities, foreseeable consequences, and foreground and background detail. It used color and more realistic rendering.

For narrative, participants selected one of ten prompts, such as describing a holiday tradition, favorite vacation, or first job, and spoke for approximately 60 seconds.

### Recording and preprocessing

Samples were collected in a quiet, private room with the examiner and participant seated across a table. A Zoom H1 Handy Recorder with fixed X/Y stereo microphones was placed approximately 12 to 18 inches from the participant. Recording was started and stopped for each prompt.

Recordings were trimmed from the end of the examiner prompt to the participant’s final utterance. Examiner speech within this window was excluded by timestamp; this occurred in four Cookie Theft recordings. Inter-word intervals spanning an excluded segment were not counted as participant pauses. Five participants stated after 50 seconds that they had nothing further to say about the Cookie Theft picture, and the recording was stopped; their shorter responses were retained. Recordings extending beyond 60 seconds because of delayed stopping were truncated at 60 seconds.

### Transcription and quality control

Recordings were transcribed with the Whisper large-v3 model [9] using the faster-whisper implementation [10], word-level timestamps, English language setting, and beam size 5. Voice activity detection was disabled to preserve silent intervals. Transcripts were reviewed against audio for recognition errors, examiner speech, and remarks unrelated to the prompt, such as comments about instructions or the recording process. Word timestamps were not manually corrected.

### Measures

Response duration spanned the first participant-word onset to the final participant-word offset; initial latency and trailing silence were excluded. A token was a transcribed word or filled pause (e.g., “um” or “uh”). Speech rate was tokens per minute of response duration; word count excluded filled pauses. Pauses were identified as onset-to-onset intervals of 2 seconds or longer, following the inter-response-interval approach used in category fluency [7]. The threshold isolated prolonged inter-word pauses rather than brief articulatory hesitations. Pause duration was measured from the preceding word offset to the following word onset. Because automatic transcription occasionally extended word offsets into silence, each effective offset was capped at 1.5 seconds after its onset. Pause time ratio was summed pause duration divided by response duration. Pause-adjusted rate was tokens per minute after subtracting detected-pause duration; shorter gaps remained included. It is not a measure of articulatory speed.

### Analysis

Cross-task correspondence was assessed using Spearman rank correlations for all task pairs and measures. Mean coefficients are arithmetic means of constituent correlations and are descriptive. No a priori power analysis or multiple-comparison correction was applied because analyses were exploratory; interpretation emphasizes effect-size magnitude and ordering. Analyses used SciPy in Python.

## Results

### Task characteristics

All participants completed all tasks. Category fluency produced shorter and slower output than the three connected-speech tasks. Mean speech rate was 64 tokens/min in category fluency (SD 36, range 15 to 121) versus 133 tokens/min across connected speech (range 71 to 193). The coefficient of variation was 0.57 for category fluency and 0.20, 0.25, and 0.22 for Cookie Theft, contemporary picture, and narrative, respectively. Mean pause time ratio was 0.39 in category fluency, with every participant showing a qualifying pause, versus 0.08 to 0.10 across connected tasks; four, two, and three participants, respectively, produced no qualifying pause.

### Correspondence among connected-speech tasks

Speech-rate correspondence was strong and largely consistent (Cookie Theft with contemporary picture, ρ = 0.79; contemporary picture with narrative, ρ = 0.78; Cookie Theft with narrative, ρ = 0.59; all *p* < 0.05; mean ρ = 0.72). Word count was similar (mean ρ = 0.61). Pause time ratio corresponded between contemporary picture and narrative (ρ = 0.67, *p* = 0.017) but not between either and Cookie Theft (ρ = 0.34 and 0.41; both nonsignificant).

### Correspondence between category fluency and connected speech

Correspondence increased from Cookie Theft to contemporary picture to narrative (Figure 2). Speech-rate coefficients were ρ = 0.36, 0.51, and 0.64, respectively; the narrative correlation was significant (*p* = 0.024). Pause time ratio followed the same ordering more steeply (ρ = 0.14, 0.45, and 0.71; narrative *p* = 0.009), as did word count (ρ = 0.29, 0.41, and 0.65). Narrative was the only connected task whose correspondence with category fluency reached significance across measures. Mean speech-rate correspondence with category fluency was ρ = 0.50 versus 0.72 among connected tasks.

**Figure 2.**
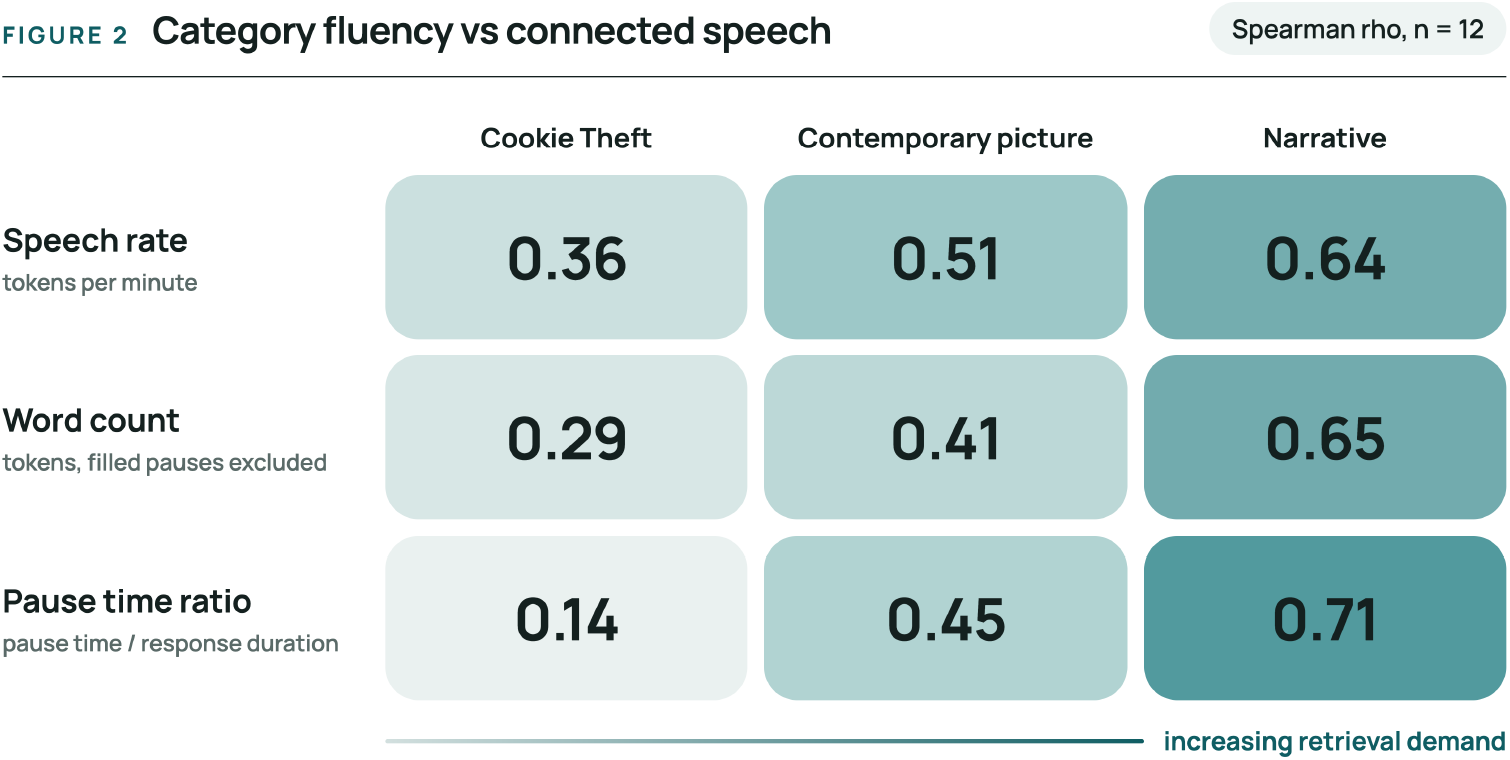
Spearman rank correlations (ρ) between category fluency and the two picture-description tasks and self-generated narrative for speech rate, word count, and pause time ratio (n = 12). Shading indicates correlation magnitude; tasks are ordered from visually constrained description to self-generated narrative.

### Individual patterns

One participant produced high-rate connected speech (131, 125, and 119 tokens/min, near the group median) but ranked third lowest in category fluency at 35 tokens/min, with a pause time ratio of 0.46. Another showed the most sustained pausing, with ratios of 0.61, 0.20, 0.22, and 0.34 in category fluency, Cookie Theft, contemporary picture, and narrative, respectively; this was the only participant elevated across all tasks.

### Pause-adjusted speech rate

This measure showed no reliable correspondence between category fluency and any connected task (ρ = 0.27 to 0.39; none significant) and an inconsistent pattern among connected tasks; it was not considered further.

## Discussion

These findings suggest that speech timing and productivity measures are not interchangeable across elicitation methods. Speech rate and word count corresponded strongly among connected-speech tasks, pause time ratio corresponded more selectively, and pause-adjusted rate showed no reliable pattern across tasks. In this sample, category fluency showed generally weaker correspondence with connected-speech tasks, reflecting greater variation in participants’ relative rankings across tasks.

Although not statistically compared, correlations with category fluency increased from Cookie Theft to contemporary picture to narrative for speech rate, word count, and pause time ratio. This pattern was steepest for pausing. One explanation could be that measures correspond across tasks to the extent that tasks share retrieval demands, with category fluency and narrative both lacking visual support despite differing Individual patterns illustrate why task selection likely matters. Some participants who were fluent in connected speech paused markedly during category fluency, with the divergence emerging only in that task. Category fluency also showed greater relative between-person variability in speech rate. This may be due to differences in the ways these tasks engage neuroanatomical networks. Category fluency tasks load more on left medial temporal structures, while connected speech tasks place broader demand on the left frontal/parietal perisylvian language network and posterior (e.g., right parietal) regions involved in visual processing. Thus differential patterns may reflect greater sensitivity to cognitive status, sensitivity to different domains of cognitive ability, or task-specific variability. Of note, category fluency is well-established as a measure sensitive to Alzheimer’s disease, whereas connected speech tasks are more specifically associated with aphasia syndromes .

Limitations include a small, highly educated sample from a single care community recorded during a single-session assessment. Analyses were exploratory and uncorrected for multiple comparisons. Available cognitive, mood, and health data were not analyzed because this study addressed cross-task correspondence rather than criterion validity. Fixed task order and variable narrative prompts limit separation of task-demand, order, and prompt effects. Automatic transcription introduced timestamp error that offset capping mitigated but did not eliminate.

Prior work establishes associations between speech rate, pausing, category fluency, and cognitive status [4,5]. This study addressed a different question: whether the same measures preserve their relative behavior across elicitation tasks. They did so inconsistently. Determining which task is most sensitive to general cognitive status requires concurrent criterion validation, but task selection should be treated as part of measurement design rather than a neutral procedural choice.

## Acknowledgements

The authors thank the participating residents and facility staff who supported this study.

## Declaration of generative AI in the manuscript preparation process

During manuscript preparation, the authors used Claude Opus 4.8 and ChatGPT to assist with language editing, clarity, and structure. The authors reviewed and verified all content and take full responsibility for the final manuscript.

## Author contributions statement

KM: Conceptualization, methodology, investigation, writing - original draft. NE: Conceptualization, investigation, writing - review and editing. TB: Conceptualization, writing - review and editing. HY: Supervision, writing - review and editing.

## Data Availability Statement

The speech recordings are not publicly available because they contain identifiable voice data from a small cohort and participant consent does not permit open sharing. De-identified measures are available from the corresponding author on reasonable request, subject to institutional approval and a data use agreement.

## Ethical Considerations

This study was approved by the Pennsylvania State University Institutional Review Board and conducted in accordance with the Declaration of Helsinki.

## Consent to Participate

Written informed consent was obtained from all participants.

## Consent for Publication

Not applicable.

## References

1. Mekulu K, Aqlan F, Yang H. A 60-second interpretable voice model for early dementia screening. PLOS Digit Health. 2026;5(7):e0001552. doi:10.1371/journal.pdig.0001552.

2. Mekulu K, Aqlan F, Yang H. A transparent four-feature speech model for depression screening applicable across clinical and community settings, including assisted-living environments. Front Digit Health. 2026;7:1675103. doi:10.3389/fdgth.2025.1675103.

3. Shankar R, Goh Z, Devi F, Xu Q. A systematic review of explainable artificial intelligence methods for speech-based cognitive decline detection. NPJ Digit Med. 2025;8(1):724. doi:10.1038/s41746-025-02105-z.

4. Cohen AS, Divers R, Calamia M, Masucci M, Granrud OE, Corporandy A, et al. Speech pause and speech rate for evaluating Alzheimer’s and mild cognitive impairment: a meta-analysis. J Int Neuropsychol Soc. 2026;32(1):24–31. doi:10.1017/S1355617725101677.

5. Konig A, Satt A, Sorin A, Hoory R, Toledo-Ronen O, Derreumaux A, et al. Automatic speech analysis for the assessment of patients with predementia and Alzheimer’s disease. Alzheimers Dement (Amst). 2015;1(1):112–124. doi:10.1016/j.dadm.2014.11.012.

6. Fraser KC, Meltzer JA, Rudzicz F. Linguistic features identify Alzheimer’s disease in narrative speech. J Alzheimers Dis. 2016;49(2):407–422. doi:10.3233/JAD-150520.

7. Troyer AK, Moscovitch M, Winocur G. Clustering and switching as two components of verbal fluency: evidence from younger and older healthy adults. Neuropsychology. 1997;11(1):138–146. doi:10.1037/0894-4105.11.1.138.

8. Clarke N, Barrick TR, Garrard P. A comparison of connected speech tasks for detecting early Alzheimer’s disease and mild cognitive impairment using natural language processing and machine learning. Front Comput Sci. 2021;3:634360. doi:10.3389/fcomp.2021.634360.

9. Radford A, Kim JW, Xu T, Brockman G, McLeavey C, Sutskever I. Robust speech recognition via large-scale weak supervision. Proc Mach Learn Res. 2023;202:28492–28518.

10. Klein G. faster-whisper: Faster Whisper transcription with CTranslate2 [computer software]. 2023. Available at: https://github.com/SYSTRAN/faster-whisper. Accessed July 20, 2026.

